# A cohort of minimally invasive spine surgeries in Nigeria

**DOI:** 10.1101/2025.03.14.25323958

**Authors:** Oluwafemi F. Owagbemi, Temitayo O. Ayantayo, Olawale A.R. Sulaiman

## Abstract

Minimally invasive spine surgery (MISS) has gained traction since its introduction into the spine surgery armamentarium, resulting in better outcomes than the traditional open approaches. It was only recently introduced in Nigeria, where it is rarely performed. In a bid to improve access to state-of-the-art neurosurgical services in his home country, the lead author, having practiced MISS in the United States, started performing it in Nigeria in 2017. We aim to describe our MISS experience in Nigeria, a lower-middle-income country (LMIC) with high poverty indices.

This is a retrospective review of our database of patients who had MISS for degenerative spine disease involving the thoracic, lumbar, and lumbosacral spine regions from April 2017 to May 2022. Demographic, perioperative, and patient-reported outcome data were retrieved and analyzed (statistical significance—p < 0.05). The same lead surgeon performed the procedures with similar operative techniques and perioperative management.

The data of the 143 patients were not normally distributed. The median age was 62 years and males comprised 55.9%. About half the patients had minimally invasive (MIS) laminectomy; 45.5% and 3.5% had MIS-transforaminal lumbar interbody fusion (TLIF) and MIS-microdiscectomy, respectively. Most (73.4%) were in the lumbar spine, 25.2% involved the lumbosacral junction, and 1.4% were in the thoracic spine. Median surgery duration, estimated blood loss, and length of hospital stay were 112 mins, 50 cubic centimeters, and 3 days, respectively. The overall perioperative complication rate was 6.3%, while surgical complications occurred in 4.9% of the patients. The patient-reported outcomes (Numeric Rating Scale [NRS] scores and Oswestry Disability Index [ODI]) showed minimal important differences (MID) between baseline and one-year follow-up.

This study’s perioperative parameters and complication (overall and specific) rates are comparable to those obtained from previous work on MIS lumbar decompression (laminectomy and microdiscectomy) and TLIF in higher-income countries. TLIF, whether open or minimally invasive, is more invasive, destructive, and technically demanding than decompression. It is, therefore, not surprising that MIS-TLIF resulted in statistically higher SDn, EBL, and LOS than MIS-decompression. The higher incidence of complications in MIS-TLIF compared with MIS-decompression is also not unexpected because of the instrumentation and implants involved in MIS-TLIF. In the same vein, it is unsurprising that patients who underwent MIS-TLIF had statistically higher discharge to inpatient physical therapy rates and lower discharge home rates than those who underwent MIS-decompression.

The improvement between median NRS and ODI scores at the preoperative evaluation and one year after surgery were either comparable to or exceeded the MID in these patient-reported outcomes demonstrated in previous research—a two-point change for NRS and a 5.9–20 difference for ODI. Our efforts to introduce MISS as part of our practice in Nigeria are informed by the need for deploying, developing, and maintaining beneficial cutting-edge care in LMICs where the capacity exists while not neglecting the ‘stock’ procedures.

MISS is available in Nigeria, and it is characterized in our practice by satisfactory patient-reported outcomes and comparable perioperative parameters and complication rates to those obtained from MISS performed in countries with higher incomes than Nigeria, where MISS is rife.

## Introduction

Low back pain (LBP) is the leading cause of disability globally, and its incidence, prevalence, and associated disability-adjusted life years (DALYs) have increased over the last two decades because of the rising life expectancy.[1–3] Lumbar degenerative disc disease (DDD) is a common cause of chronic LBP in adults,[4] while its less common counterpart, thoracic DDD, is potentially more pernicious because of the narrower thoracic spinal canal compared with the lumbar spine and the risk of thoracic myelopathy.[5] DDD, in general, is a leading disorder necessitating care in Nigeria and globally.[4,6–9]

The surgical treatment of thoracic and lumbar DDD has evolved over the years from decompression to instrumented fusion of various kinds, with minimally invasive techniques recently added to the surgical repertoire.[10,11] Minimally invasive spine surgery (MISS) has gained traction since its introduction into the spine surgery armamentarium.[12] It is designed to result in reduced collateral tissue damage and morbidity, and a faster functional recovery compared to the traditional open approaches, all with the same surgical goal.[13] True to design, the tissue damage that results from open posterior spine approaches, particularly in the multifidus muscle,[14–17] is statistically lower in MISS.[18] Surgery duration, estimated blood loss, length of hospital stay, and complication rate are also reduced in comparison.[19–22]

Given this improved outcome, it is no wonder that MISS is increasingly employed in many parts of the world,[23,24] despite its steep learning curve[25–27] and the need for more advanced equipment than required for the open approaches.[28,29] The story appears different in some regions, however. In a global survey of spine surgeons, only half (the lowest proportion in the study) of the 16 respondents in the Africa and Middle East region thought that MISS was considered mainstream in their areas and practice settings,[30] while its utilization for fusion in the region was one of the lowest in another worldwide survey.[31]

MISS has been reported for the treatment of spine trauma, tuberculosis, and degenerative disease in Tanzania and South Africa (both African countries),[32–34] and has also been used in Haiti.[35] Tanzania and Haiti, like Nigeria, are classified as lower-middle-income economies, while South Africa is an upper-middle-income economy.[36] We did not find a report of MISS from Nigeria, where, based on anecdata, it is both new and rare. In the bid to improve access to state-of-the-art neurosurgical services in his home country, the lead author (OARS), having practiced MISS in the United States (U.S.), started performing it in Nigeria in 2017. This study aims to describe the authors’ MISS experience in Nigeria and present the associated perioperative parameters, complications, and patient-reported outcomes.

## Materials and methods

The lead author maintained a prospective database of the patients in his practice at RNZ Clinic/Euracare Multi-Specialist Hospital, Victoria Island, Lagos, Nigeria. The data includes patient demographics, diagnoses, and perioperative parameters, including the surgeries performed, the number of spinal levels (NOL) operated on, surgery duration (SDn), estimated blood loss (EBL), length of hospital stay (LOS), perioperative complications, patient-reported outcomes (PRO), and discharge destination. For this study, we performed a retrospective review of the listed data in patients who had minimally invasive spine procedures for degenerative spine disease (DSD) involving the thoracic, lumbar, and lumbosacral spine regions from April 2017 to May 2022. The database was accessed for the research on January 31, 2024, and only one author, one of the physicians in the practice, had access to information that could identify individual participants during data collection. Patient consent was obtained for all procedures, and the Nigerian Institute of Medical Research’s Institutional Review Board granted ethical approval for the study.

We made the diagnosis of thoracic, lumbar, and lumbosacral DSD from a combination of clinical features of mechanical back pain, radiculopathy, and/or neurogenic claudication, with magnetic resonance imaging findings of spondylosis, spondylolisthesis, spinal canal stenosis, and/or neural foraminal stenosis. Following six weeks of unsuccessful non-operative treatment, we recommended surgery. SDn and EBL were determined from operating room records and LOS from admission records.

The perioperative complications that were considered for the study are dura tear, early surgical site infection (based on the Centers for Disease Control and Prevention, USA criteria and within 30 days of surgery),[37] malpositioned implants, venous thromboembolism, and nerve injury. A dura tear was recorded when observed during surgery or if postoperative cerebrospinal fluid (CSF) leakage was observed. We evaluated the surgical wounds per practice protocol—as needed during admission, on the seventh postoperative day, and one month after surgery, with instructions for features of wound infection given to the patient and caregivers at discharge. Malpositioned implants were diagnosed using postoperative computed tomography confirmation with or without dermatomal pain. Pulmonary embolism (PE) diagnosis was made using clinical and electrocardiogram parameters, with a computed tomography pulmonary angiogram, when the patient’s poor clinical status did not obviate one. Nerve injury was recorded when observed during the surgery and when the patient developed new weakness after surgery.

The patient-reported outcomes that we evaluated were low back pain and leg pain, using the 11-point (from 0 for no pain to 10 for the worst imaginable pain) Numeric Rating Scale (NRS),[38] and disability, using the Oswestry Disability Index (ODI).[39,40] These were assessed preoperatively and at six weeks, six months, and one year after surgery, using questionnaires. Apart from surgical site infection (SSI) and the PROs, observation of the variables, because of their nature, was limited to the admission period.

The lead author performed all the minimally invasive (MIS) procedures—minimally invasive transforaminal lumbar interbody fusion (MIS-TLIF), minimally invasive laminectomy (MIS-laminectomy), and minimally invasive microdiscectomy (MIS- microdiscectomy)—using a similar technique to that employed in a previous publication,[19] assisted in some by the co-authors. MIS-laminectomy and MIS- microdiscectomy were grouped into minimally invasive decompression (MIS- decompression) for ease of analysis. We irrigated all wounds copiously and closed them in layers. All the patients had similar postoperative care using standardized postoperative order sets, and the care for each procedure type was nearly identical for the patients who underwent it. Patients were sent for physical therapy two to four days after surgery as needed, and they returned to work as soon as they could. The postoperative management is also similar to the lead author’s method in the U.S.[19]

### Statistical analysis and study reporting

The patients’ demographics, DDD data, and surgery types were the independent variables, while SDn, EBL, LOS, complication rates, disposition data, and PROs constituted the dependent variables. The data were evaluated using the Shapiro-Wilk test for normality, to determine whether to use parametric and nonparametric tests, and described using counts, percentages, and medians. We used the two-tailed Mann-Whitney U test to compare variables with nonparametric continuous data, while for categorical data, the chi-squared test was used to evaluate associations and goodness of fit. Where the conditions of the chi-squared test were not met, Fisher’s exact test was used. Non-parametric repeated measures analysis of variance was used to compare the baseline patient-reported outcomes with the follow-up values. We set statistical significance at p < 0.05[41,42] and missing data were addressed using the available-case analysis.[43] We performed the statistical analysis with *jamovi* (The jamovi project, Sydney, Australia)[44] and implemented the Strengthening the Reporting of Observational Studies in Epidemiology (STROBE) guideline in reporting this work.[45]

### Study environment

Nigeria had a low Human Development Index (HDI) of 0.535 and ranked 163 of 191 nations in the 2021/22 United Nations Development Programme Human Development Report.[46,47] Lagos, the state in Nigeria where our practice is located, was reported over multiple years to have the highest HDI in the country.[47,48] With an HDI of 0.681 in the 2021/22 report, which is better than the national HDI, Lagos falls into the medium human development range, but lags behind Western countries with very high development indices, and even some African countries in the high development range.[46,47] When told through another metric, the World Bank’s Human Capital Index (HCI) of 2020, which scores the country an index of 0.36 (the HCI ranges between 0 and 1), Nigeria’s story remains one of poor development.[49]

The underdevelopment translates into the people’s pockets, affecting their purchasing power and healthcare. With a population of about 220 million people,[50] Nigeria has 40.1% of its population living below the national poverty line and 39.1% living below the international poverty line of $1.90 a day.[46] This is further worsened by poor healthcare access that is connected to poor social health insurance coverage, which was recently reported to be less than 5% of the population, with most people paying out-of-pocket for healthcare.[51] This out-of-pocket health expenditure contributes to impoverishment in the country.[52]

On the backdrop of this socioeconomic scenario, and even with neurological diseases and injuries being major causes of death and disability globally,[53,54] neurosurgery suffers healthcare disparities between developed and developing countries, with low-income countries (LIC) and lower-middle-income countries (including Nigeria) having poor access to neurosurgical care.[55] Degenerative spine disease is one of the leading conditions requiring such care globally, including in Nigeria.[4,6–9] Despite the stark realities of healthcare in Nigeria, efforts to provide safe and affordable neurosurgical care in the country are being made, especially by individuals and through private institutions, including the lead author, who moved his practice completely from the U.S. to Nigeria in 2019.[56]

This practice is in a private hospital with a single operating room equipped with a Jackson table, Wilson frame, operating microscope, and fluoroscopy machine, which were used as standard items for MISS. Retraction was maintained during the MISS procedures using a 22 mm tubular retractor (METRx System, Medtronic, Memphis, TN, USA), while pedicle screw instrumentation was performed percutaneously. Some of the other neurosurgical conditions treated in the practice include brain and spine tumors, chronic subdural hematomas, brain and spine trauma, and peripheral nerve injuries and entrapments.

## Results

A total of 261 patients had spine surgery during the 62-month study period, 143 of whom had minimally invasive spine procedures for DSD involving the thoracic, lumbar, and sacral spine regions. This constitutes 54.8% of our spine surgery practice and 65.0% of the 220 patients operated on for DSD during the period. The excluded patients had open spine surgeries and/or surgery for non-degenerative spine conditions. When tested for normality, the data were not normally distributed.

The median age was 62 years, with an interquartile range (IQR) of 15 years, and males comprised 55.9% (80/143). About half (73, 51.0%) of the patients had MIS- laminectomy, while 65 (45.5%) and five (3.5%) patients had MIS-TLIF and MIS- microdiscectomy, respectively. Overall, 105 (73.4%) of the surgeries were performed in the lumbar spine, while 36 (25.2%) involved the lumbosacral junction and the remaining two were in the thoracic spine (Fig 1). All the MISS were performed in one (82, 57.3%) or two (61, 42.7%) spinal levels.

**Fig 1.**
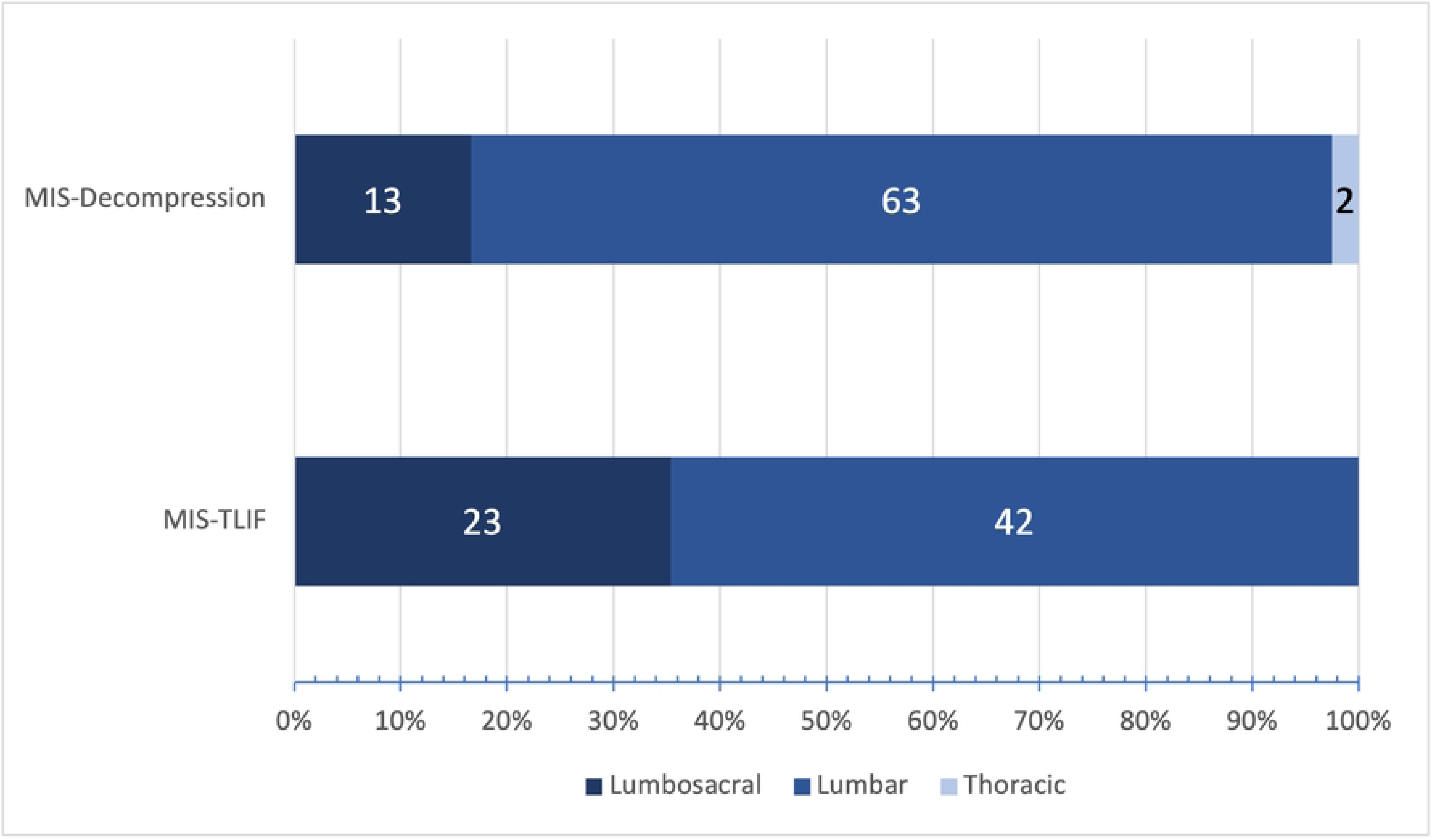
Spinal regions operated on—comparison of minimally invasive transforaminal interbody fusion (MIS-TLIF) with minimally invasive decompression (MIS-decompression), which comprises minimally invasive laminectomy and minimally invasive microdiscectomy.

The median (IQR) SDn, EBL, and LOS were 112 (121) min, 50 (70) cubic centimeters (cc), and 3 (0) days, respectively. There were three dura tears (2.1%), two incidents each of PE and malpositioned screws (1.4% each), and one SSI and nerve injury each (0.7% each), with complications directly related to surgery occurring in 4.9% and an overall complication rate of 6.3%. A subanalysis of the patients into MIS-TLIF and the MIS-decompression is shown in the table. With age (p = 0.334) and NOL (p = 0.354) operated on being similar in both groups (see Table), MIS-TLIF had higher SDn, EBL, and LOS values (all p < 0.001), and complication rate (p = 0.011).

**Table.**
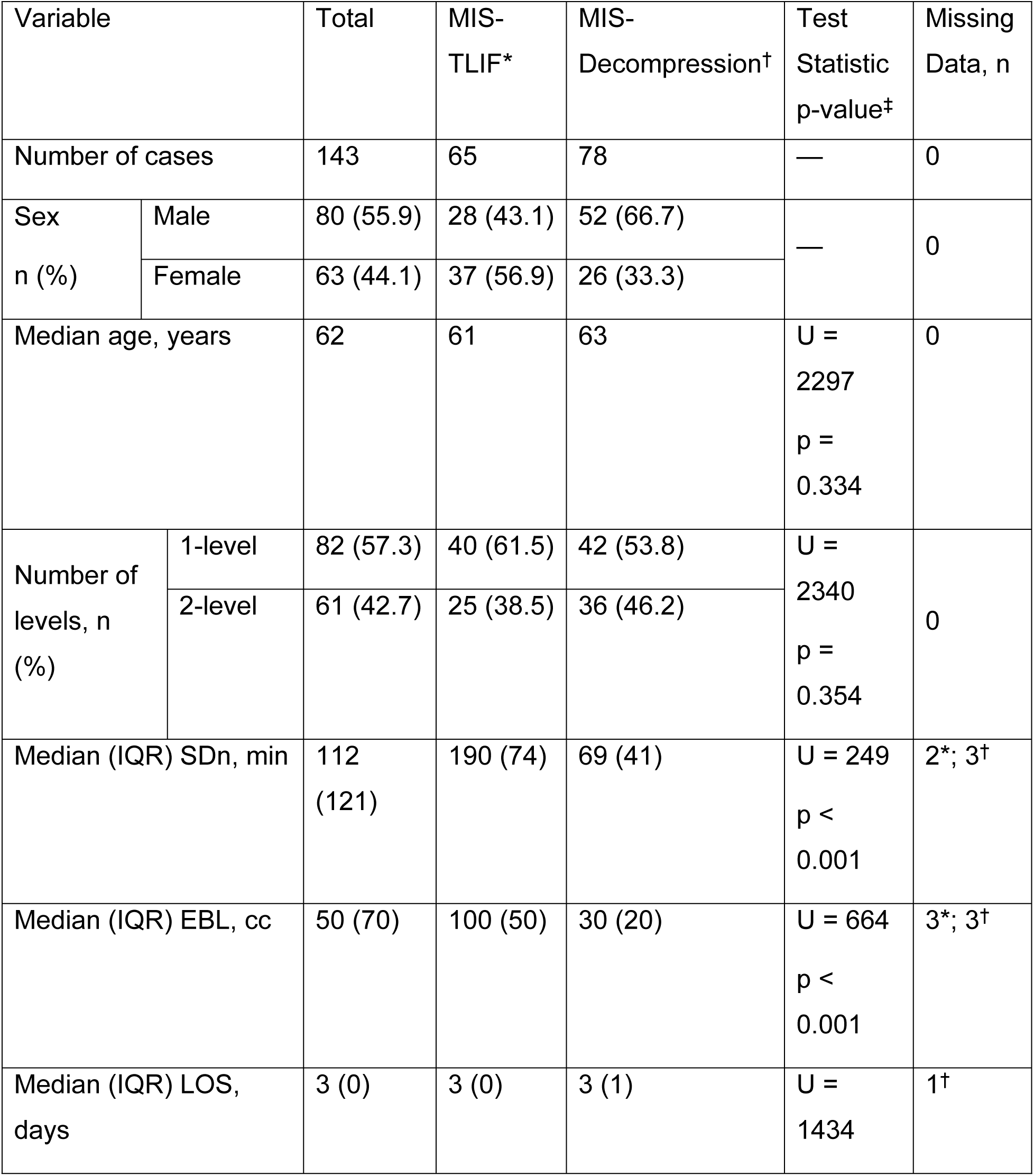
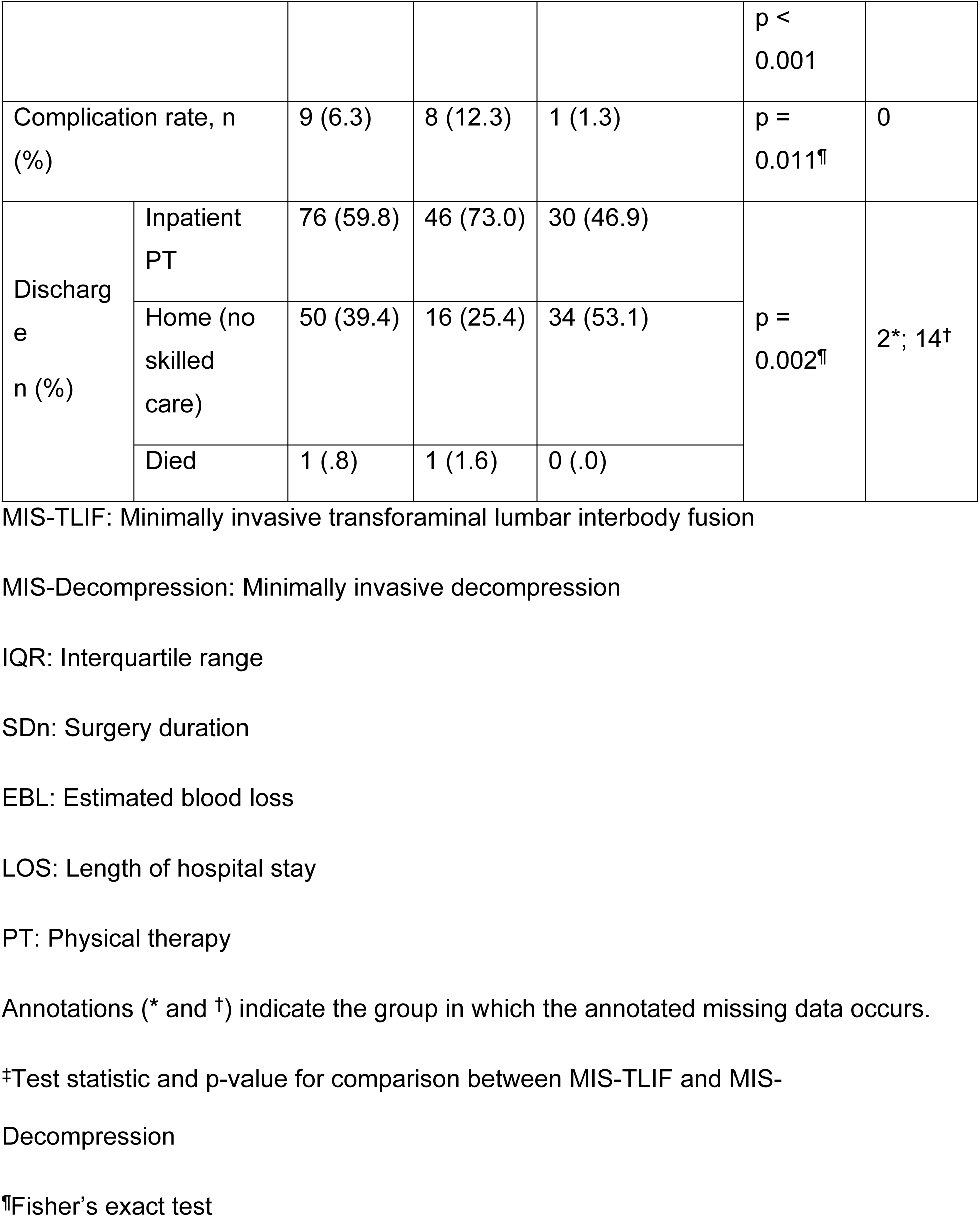
Comparison of minimally invasive transforaminal interbody fusion with minimally invasive decompression, which comprises minimally invasive laminectomy and minimally invasive microdiscectomy.

Each malpositioned screw required an additional procedure for revision in the two affected patients. The patient who developed SSI had undergone MIS-TLIF and was one of those who had a dura tear, which led to postoperative CSF leakage, for which he had a second procedure for dura repair. The SSI was a deep one, which caused prolonged admission and required an additional procedure for wound debridement and washout. The nerve injury, with an associated dura tear, occurred intraoperatively during MIS-TLIF, and this resulted in a right-sided foot drop, which improved with physiotherapy. The third dura tear occurred during an MIS-laminectomy, being the only complication in that group. The latter two dura tears were repaired in the same surgery with no postoperative CSF leakage. The two patients who suffered PE had undergone MIS-TLIF, and one of them was a morbidly obese woman who died on the seventh postoperative day after developing sudden severe PE symptoms and rapid clinical deterioration. The other patient’s symptoms were insidious, and she did well with anticoagulation therapy. There were no complications in the patients who had MIS-microdiscectomy.

Most (76, 59.8%) of the patients were discharged to inpatient physical therapy, while 39.4% (50) were discharged home with no skilled care (see Table for comparison between MIS-TLIF and MIS-decompression). A statistically higher proportion of patients less than 65 years of age were discharged home compared to patients greater than/equal to 65 years of age (p = .01).

The median NRS score was 8.0 preoperatively, while postoperatively, the median scores were 4.0, 5.0, and 3.5 at six weeks, six months, and one year, respectively. There was a statistical difference between the baseline NRS score and the follow-up values (each p < 0.001) during pair-wise comparison. The preoperative median ODI was 56%, while it was 38%, 36%, and 26%, respectively, six weeks, six months, and one year after surgery. There was also a statistical difference between the baseline ODI and the follow-up values (p = 0.048, p = 0.008, and p < 0.001, respectively) during pairwise comparison. The NRS and ODI results are further illustrated in Figs 2 and 3.

**Fig 2.**
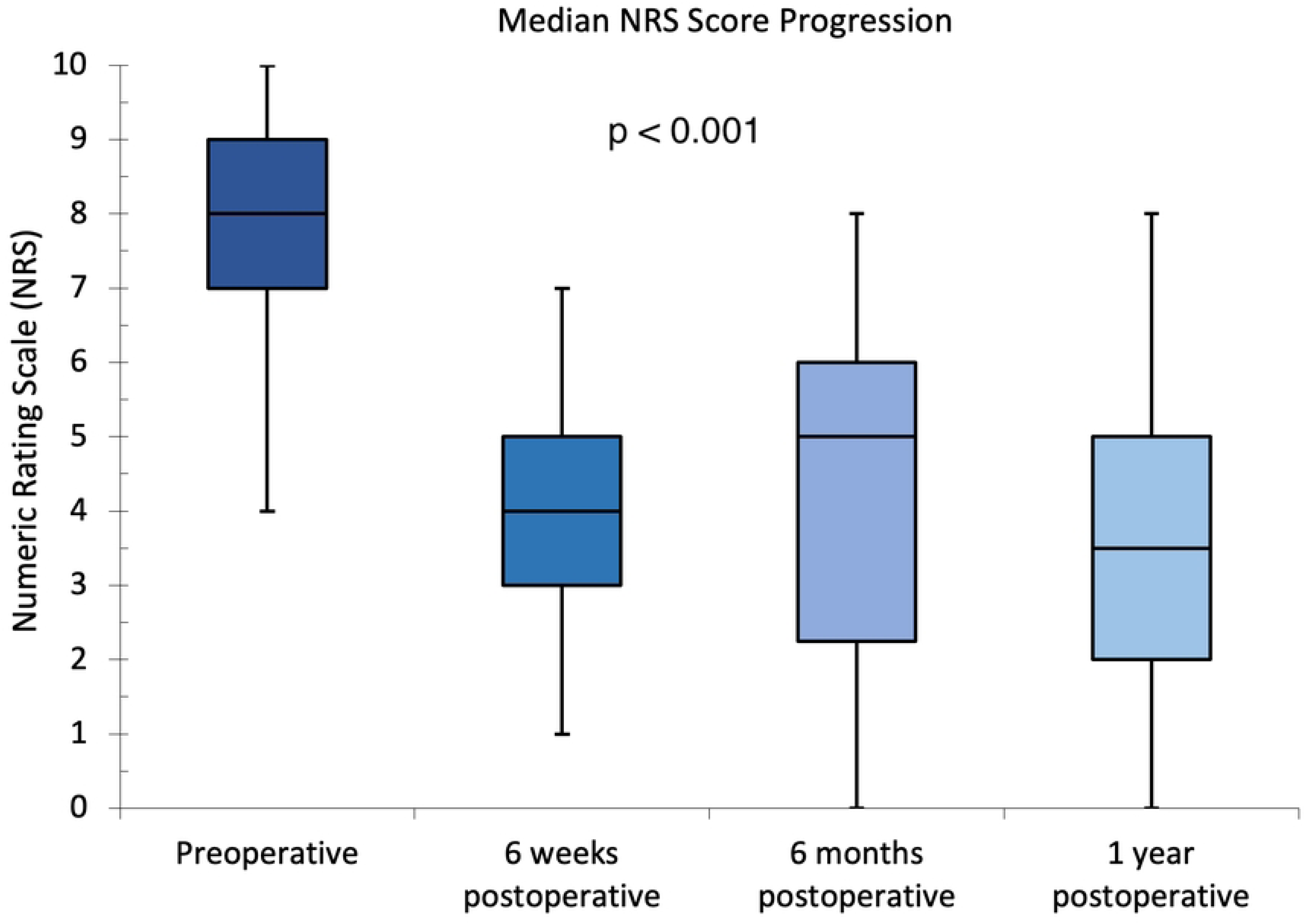
Median Numeric Rating Scale scores at preoperative and follow-up evaluations.

**Fig 3.**
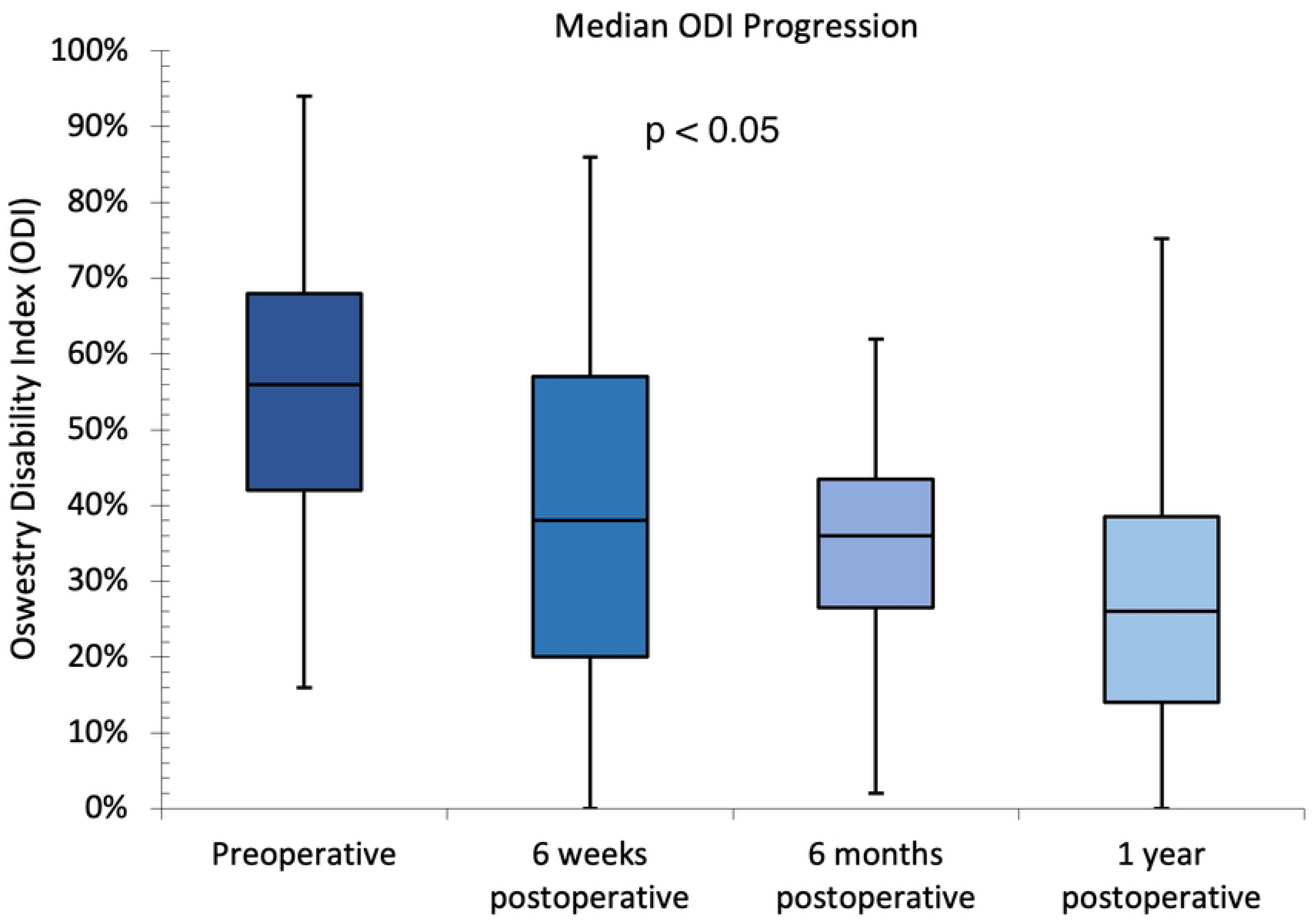
Median Oswestry Disability Index at preoperative and follow-up evaluations.

## Discussion

The results of this study describe the patients, indications, practice, perioperative parameters, complications, and patient-reported outcomes of MISS performed by the authors in Nigeria. Though the data were prospectively collected, some were missing. They were assumed to be missing completely at random (MCAR) since they were not observed to be related to other variables.[57] We believe that applying the available-case analysis to the missing data helped keep the data as robust as possible.[43] These missing data are likely due to omissions in records.

This study’s perioperative parameters and complication (overall and specific) rates are comparable to those obtained from previous work on MIS lumbar decompression and MIS-TLIF.[19,58–69] While there is much similarity in surgical techniques and outcome evaluation between these reports and our work, there are a few differences we wish to highlight. In contrast to this study and the work of Sulaiman et al.,[19] where bilateral percutaneous screw fixation was performed, Nandyala et al.[59] used a unilateral approach, which may explain the lower SDn and EBL they found, particularly in the second arm of their study. Another difference is the much higher complication rates of Ahn et al.[58] and Nandyala et al.[59] This is likely due to their inclusion of longer-term complications, like pseudoarthrosis, which we did not set out to evaluate.

Patients’ home discharge rates vary across studies, given that the patient populations and techniques in these studies differ.[65–69] Our patient disposition data compare well with those from a “spine naïve” community hospital in the U.S., also reporting initial cases.[66] The dearth of skilled home care in our environment markedly hampered our ability to discharge patients needing skilled care to their homes. Our finding that the discharge to inpatient physical therapy rate was statistically higher for older patients compares with the work of Altshuler et al.,[69] though they used 60 years as their threshold. Most of the patients with missing disposition data in our study were less than 65 years of age. A mock analysis of a scenario where all the patients below 65 years of age who had missing data were discharged home still yielded a statistically higher inpatient physical therapy discharge rate for older patients.

TLIF, whether open or minimally invasive, is more invasive, destructive, and technically demanding than laminectomy and microdiscectomy. It is, therefore, not surprising that MIS-TLIF resulted in statistically higher SDn, EBL, and LOS than the MIS-decompressions as also observed by Chan et al.[65] in their study. In the same vein, it is also not surprising that patients who underwent MIS-TLIF had statistically higher discharge to inpatient physical therapy rates and lower discharge home rates than those who had MIS-decompression. The higher incidence of complications in MIS-TLIF compared with MIS-decompressions is also not unexpected because the instrumentation involved in MIS-TLIF increases the risk of dura and nerve injuries. Furthermore, implants increase SSI risk and may be misplaced intraoperatively.[70] The only SSI in this study occurred in a patient who had MIS-TLIF. Though both PEs occurred in MIS-TLIF patients, there is no indication that MIS-TLIF is an independent risk factor for PE.

Patient-reported outcomes are important and are increasingly used in spine surgery, particularly regarding minimal important difference (MID), which is the smallest change perceived by patients as clinically important.[71,72] In our study, the improvement between median scores at the preoperative evaluation and one year after surgery were either comparable to or exceeded MIDs found by other researchers—a two-point change for NRS and 5.9–20 for ODI[72]—and the difference was statistical.

Improved access to safe and affordable surgical care is both a major need in lower-middle-income countries (LMIC) and a 2030 Global Surgery target.[73] The healthcare disparities between developed and developing nations that affect neurosurgery[55] will also need to be addressed if this need/target must be met. The neurosurgical workforce is an area of disparity, with few neurosurgeons dealing with huge loads of essential consultations and surgical cases in these developing countries, and the workforce need is projected to worsen towards 2030.[55,74] While capacity-building efforts like twinning and training are helpful, these methods face challenges,[75] some of which are likely to be mitigated by the continuous presence of the capacity builders in the countries of need, as is the case in our practice. The benefit is seen in neurosurgical care generally, but where the capacity for beneficial cutting-edge care can be deployed, developed, and maintained in LICs and LMICs, we believe it is expedient to offer such care to the people in these nations, while not neglecting the ‘stock’ procedures. This informed our efforts to introduce MISS as part of our practice in Nigeria.

Despite the advantages of MISS (reduced complication profile and increased safety compared to open approaches), its resource-intensiveness has made it generally prohibitive in developing countries, leaving its availability there to the episodic interventions of practitioners from countries in the Global North.[32,35,76] The pitfalls of this approach, despite the good intentions, possibilities, and successes, have been described.[76] The feasibility of MISS as a continuous practice in the Global South rather than as isolated occurrences[32–35] is shown in this study. Though it started as monthly visits, the lead author’s MISS practice in Nigeria has morphed into a steady practice, with patient numbers approaching those of his previous practice in the U.S. If there are reports of more consistent MISS practice in the region, we have yet to find such, despite the reported acceptance and utilization in the Africa and Middle East region in the cited global surveys.[30,31] Since both ‘subregions’ were combined in the surveys, perhaps the acceptance and utilization were recorded more in the Middle East than in Africa.

Our study’s limitations include its retrospective nature with its inherent challenges and the associated missing data that has been discussed. In addition, it is a single surgeon and single center experience, which is not implausible since MISS is just burgeoning in Nigeria. Nonetheless, we believe it is important to highlight this work to explore the possibilities of MISS in the country. As the practice of MISS spreads in Nigeria, multicenter studies may corroborate our findings, making our results generalizable. One of our study’s strengths is that the patient numbers approach those of multiple studies from the developed world over similar periods. There is also strength in the evaluation of the parameters using sources free of surgeon bias, such as anesthesia and nursing records, as well as patient-filled questionnaires.

## Conclusions

MISS is available in Nigeria, and it is characterized in our practice by satisfactory patient-reported outcomes and comparable perioperative parameters and complication rates to those obtained from MISS performed in countries with higher incomes than Nigeria, where MISS is a staple of spine surgery practice. This underscores the role that diaspora medical experts can play in the development of modern surgical care in Sub-Saharan African countries. A reversal of the “brain drain” to “brain gain” is slowly happening in many medical fields in Nigeria, whereby U.S.- and U.K.-trained specialists are returning to Nigeria to provide state-of-the-art healthcare to Nigerian patients.

## Data Availability

An accession number and/or DOI will be made available after acceptance.

